# Safe Autopsy Procedures for COVID-19: Experience of One Research Center

**DOI:** 10.1101/2025.05.01.25326750

**Authors:** Madison P. Cline, Analisa M. Stewart, Anthony J. Intorcia, Jessica E. Walker, Courtney M. Nelson, Richard A. Arce, Michael J. Glass, Daisy Vargas, Sanaria H. Qiji, Claryssa I. Borja, Spencer J. Hemmingsen, Addison N. Krupp, Rylee D. McHattie, Kayleigh N. Martinez, Jaclyn Papa, Javon C. Oliver, Aryck Russel, Katsuko E. Suszczewicz, Holly M. Hobgood, Monica Mariner, Ileana Lorenzini, Sidra Aslam, Cecilia Tremblay, Suet Theng Beh, Lucia N. Sue, Thomas G. Beach, Geidy E. Serrano

**Author notes:** Corresponding author: Geidy E. Serrano, PhD Civin Laboratory for Neuropathology Banner Sun Health Research Institute 10515 W Santa Fe Drive Sun City, AZ 85351.

## Abstract

In the early days of the 2019 coronavirus disease (COVID-19) pandemic, there were few published guidelines for safely performing autopsies of infected individuals. The limited information relied on assumptions that the risks would be similar to those of other primarily respiratory infections, but there remained a considerable number of uncertainties, particularly in regard to the potential for aerosol transmission. Despite these novel risks, it was crucial for our program to quickly gain more knowledge about this new pathogen by continuing to perform autopsies while concurrently mitigating the risk of infection to autopsy personnel. Between January 26, 2020, the date of the first confirmed COVID-19 case in Arizona, and February 28, 2022, the Arizona Study of Aging and Neurodegenerative Disorders and its Brain and Body Donation Program performed autopsies on 162 subjects, of which 36 tested positive for SARS-CoV-2 on FDA-certified PCR tests while 2 were undiagnosed. In response to the increasing case rate in Arizona, major changes to the autopsy protocol were initiated in mid-March 2020, piloted by the autopsy director. While the new protocols were evolving and being implemented, autopsies were restricted to the brain in order to eliminate exposure to highly infectious respiratory tract tissues. Whole-body autopsies were resumed on June 6, 2020, after implementation of new protocols, which included engineering, environmental, and procedural changes designed to limit the risk to autopsy personnel. Due to a reasonable assumption that aerosol transmission was possible or probable, a key protocol change was our usage of a simple method for containment of bone saw-generated aerosols during skullcap removal, by using a heavy-duty clear polyethylene bag sealed at the subject’s neck and saw handle. Spinal cord removal was permanently suspended due to the challenges of containing the aerosols. During this reported time period, there were 94 COVID-19 autopsy exposures involving 19 staff members, with only 2 occurrences of a single autopsy team member testing positive for SARS-CoV-2, by an FDA-licensed PCR test, within 7 days of their participation in a SARS-CoV-2-positive autopsy. During this same time period, 9 autopsy personnel tested positive without this temporal proximity to autopsy participation. This suggests that our revised protocols, when adequately implemented, drastically reduces any enhanced risk of SARS-CoV-2 infection to autopsy personnel beyond the normal risk already established within the community. We therefore recommend the use of our protocol to other laboratories performing autopsies on subjects with health-threatening communicable conditions.

## Introduction

In the early days of the 2019 coronavirus disease (COVID-19) pandemic, there were few published guidelines for autopsies of infected individuals, and these were based only on assumptions that the risks would be similar to those of other primarily respiratory infections. There remained a considerable number of uncertainties, particularly about the potential for aerosol transmission and the level of respiratory protection needed (Ha JF, 2020). Due to these uncertainties and the rapid spread and lethality of SARS-CoV-2, autopsy examinations on cases of COVID-19 were completely suspended at many medical centers (Loibner et al, 2020; Salerno M et al, 2020; Akinyemi et al, 2023) and SARS-CoV-2 studies in human postmortem organs were delayed.

Autopsy remains the gold standard for pathologic investigation and provides information about the way disease is expressed in the human body that cannot be obtained through any other means.

Autopsies often provide insights that clinical studies alone may have missed. They also enable the validation of clinical findings through correlative clinicopathological studies and are important to validate and help the development of better clinical diagnostic markers and treatments. Autopsy studies done in the first months of the COVID-19 pandemic provided a critical understanding of the full effects of SARS-CoV-2 infection (Aquila et al, 2020; Carpenito, L et al, 2020; Skok, Kristijan et al, 2021; Bryce et al, 2021) and therefore it was crucial to continue these albeit with revised protocols that would allow optimal protection to exposed autopsy personnel.

The Royal College of Pathologists was among the first to offer COVID-19 autopsy guidelines for clinical and research practice. These guidelines gave recommendations for appropriate PPE to be worn, mortuary factors to be taken into consideration, as well as pathological findings to look for with a COVID-19 infection (Hanley et al, 2020 and Osborn et al 2020). Different levels of PPE were and are still recommended depending on the degree of interaction with a COVID-19 infected individual. These recommendations varied based on who set forth the guidelines and could vary greatly between countries (Baj et al, 2021). The general consensus for PPE needed for handling a body infected with COVID-19 in the United States was to use a disposable gown, hand hygiene, a waterproof apron, facemask or goggles, double gloves, N-95 respirators, shoe caps, and hair nets (Dijkhuizen et al, 2020). These recommendations were largely based on assumptions, as at the beginning of the COVID-19 pandemic, little was known about the transmission rate. It was also unknown if an individual who died with an active Sars-Cov-2 infection remained infectious postmortem. As the pandemic progressed, new research showed that the virus was still present in the body even up to a month after death (Dell’Aquila et al, 2020; Plenzig et al, 2021; Sablone et al, 2021).

Autopsy personnel have the potential to have laboratory acquired infections by contact with droplets, direct cutaneous inoculation, and aerosol exposure, and as many infections are undiagnosed before autopsy, there is always an unknown level of risk (Nolte et al, 2002). Aerosols are known to be possible transmission routes for different pathogens such as hepatitis viruses, Human Immunodeficiency Virus, and Mycobacterium tuberculosis, all of which have been known to cause infections after autopsy exposure (Burton JL, 2003; Droge et al, 2022; Johnson et al 1991; Pluim et al, 2019; Nolte et al, 2002).

Brain removals were suspected to be particularly hazardous in COVID-19 due to the aerosols generated by powered bone saws and thus were recommended to be restricted or done only with manual saws, vacuum shrouds or physical barriers (Briefing from Royal College of Pathology, 2020; Centers for Disease Control, 2020; Hanley et al, 2020, Aquila I, 2020). Oscillating saws create a cloud of particles around the operator, who therefore needs special protection (Hamilton et al, 2023; Pereira et al, 2012; Kernbach-Wighton et al, 1996; Wenner et al, 2017).

Aerosols that are smaller than 5 μm are known to stay in the air for long periods of time and are the most dangerous as they are within the respirable range (Pluim et al, 2018; Jones et al, 2015; Nolte, 2002). Various methods have been used to reduce aerosol-generating-procedures (AGPs), including using hand saws, blades, or shears as opposed to oscillating saws, or vacuum suctions attached to oscillating saws to collect bone dust (Dijkhuizen et al, 2020; Skok et al, 2021; Pluim et al, 2018; Droge et al, 2022). More elaborate techniques have included custom-built craniotomy boxes (Cheshire et al, 2021; Thakral et al, 2023; Hasmi et al, 2020). In addition, ventilation systems with HEPA filters, as well as Powered Air Purifying Respirators (PAPR) can be used to help protect autopsy staff (Pluim et al, 2018; Wenner et al, 2017). Testing of commercially available PAPRs have shown that the concentration of particles was 1.8 x 10^5^ lower inside the PAPR while sawing a calf bone, whereas unprotected, a person would generally inhale particles from an oscillating autopsy saw at a rate of 2.43 x 10^4^ particles per minute (Wenner, et al, 2017).

The technique utilized by our Brain and Body Donation Program (BBDP) autopsy team was simple and cost effective. It was first proposed by MacArthur and Schneiderman (1986) to address the AIDS epidemic and involved using a clear plastic bag to enclose the head and saw to contain aerosols. Towfighi et al (1989) further elaborated this with rigid supports into a tent-like structure. We adopted, with a slight modification, the simpler approach of MacArthur and Schneiderman. We eliminated spinal cord removal due to the challenges of containing saw aerosols over the length of the body. Otherwise, we reduced organ sampling and handling and implemented personal protective equipment (PPE) as well as engineering, environmental, and procedural changes essentially as recommended by the Royal College and CDC. For respiratory protection we elected to go beyond those recommendations and adopt the use of PAPRs at all autopsies. We believe that what we learned from our COVID-19 experience (Serrano et al, 2022) will be useful to autopsy programs in the event of future infectious disease outbreaks.

## Methods

### Research subjects

The Brain and Body Donation Program (BBDP) is a part of the Arizona Study of Aging and Neurodegenerative Disorders, a longitudinal clinicopathological study conducted at Banner Sun Health Research Institute in Sun City, Arizona, USA (Beach et al, 2015). The Program has been in operation since 1987, and has performed over 2,200 autopsies, specializing in rapid autopsies of volunteer research subjects with a median post-mortem interval (PMI) under 4 hours. These individuals receive assessments of medical, neuropsychological, and movement disorders while alive and then have complete pathological examinations of both brain and bodily organs after death (Beach et al, 2015).

This report is focused on safety procedures instituted for COVID-19; for details of our scientific studies of COVID-19, see Serrano et al (2022).

### Snapshot of autopsy protocol before the Covid pandemic

In the pre-COVID-19 Era, autopsies at BBDP were conducted with an average of four people present. One individual focused on removing the brain (brain team leader), and one focused on removing the body organs (body team leader), both with a dedicated helper. The program also often hosted visiting researchers and students interested in observing autopsies; it was not uncommon for up to ten people to be present at a given autopsy. The dedicated morgue is a room with air exhaust to the roof, far from any pedestrian areas, with an area of 715 sq. ft. At autopsy, personnel donned and doffed within the autopsy room. Personnel wore scrubs, fluid resistant or impermeable gowns along with aprons, sleeve guards, face shields, shoe covers, surgical head coverings, surgical masks, and sterile gloves. During the procedure, organs were taken out of the body, measured, and weighed before samples being taken and placed on dry ice or in neutral buffered formalin (NBF). Brains were removed with an oscillating saw and sliced coronally, with the right hemisphere sliced frozen on dry ice and the left hemisphere slices fixed in NBF. Spinal cords were removed after exposure with an oscillating saw.

### Engineering changes instituted for COVID-19

In order to better protect autopsy personnel, engineering and environmental changes were made according to Royal College and CDC recommendations. These included the designation of a dedicated donning room and construction of a dedicated doffing room. The new doffing room was built adjacent to the morgue, opposite to the morgue entrance, and, as with the morgue itself, had negative air pressure relative to surrounding rooms as well as HEPA air filtration. The air handler exchange is 27 air changes per hour (ACH) and utilizes a MERV-11 filter. All materials necessary to complete the autopsy were stocked in the suite prior to beginning, allowing the doors to remain closed for maintaining negative air pressure in the room. Infrequently used supplies were moved out of the morgue/autopsy suite to reduce the number of surfaces present.

### PPE and donning and doffing changes instituted for COVID-19

Changes to required PPE were essentially as recommended by the Royal College and CDC except for where we exceeded those guidelines (Table 1).

**Table 1.**
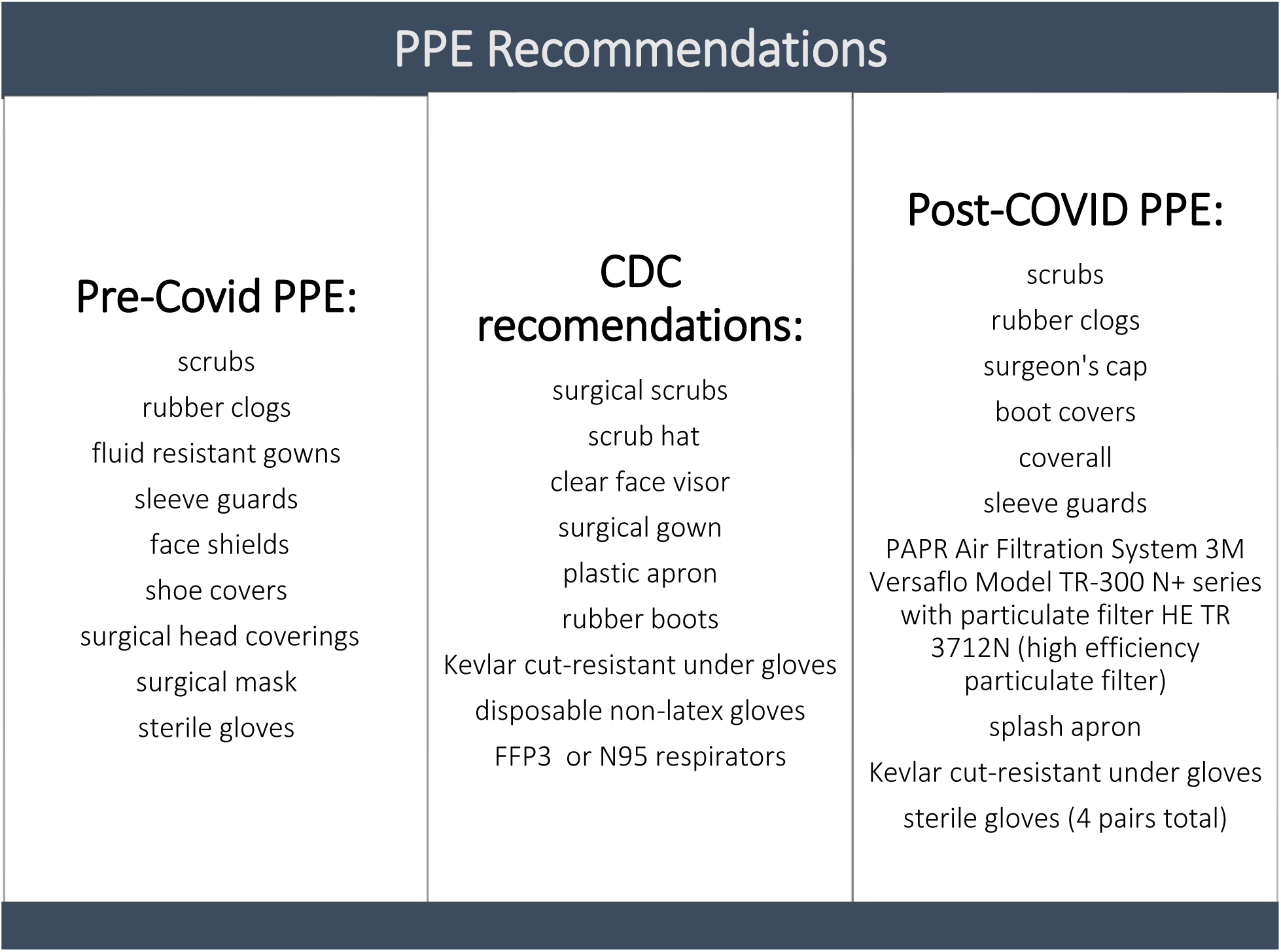
Pre- and post-COVID-19 personal protective equipment protocol at BBDP, compared with CDC COVID-19 recommendations.

Donning and doffing procedures were adopted from those designed for Ebola and other viral hemorrhagic fevers (Centers for Disease Control and Prevention, 2020). The new donning procedure required each technician to change into rubber shoes, perform hand hygiene prior to donning, and wear an N-95 mask as they don: an initial pair of disposable gloves, disposable boot covers, disposable bouffant or surgeon’s cap to cover hair and ears, a second pair of disposable gloves, puncture-resistant disposable coverall, disposable sleeve guards, a third pair of disposable gloves with gauntlet, disposable splash apron, re-usable PAPR Air Filtration System (used for all autopsies after 09/08/2020; 3M Versaflo Model TR-300 N+ series with high efficiency particulate filter HE TR 3712N), cut-proof Kevlar glove on non-dominant hand, and a fourth pair of disposable gloves.

While donning, partners observe each other, inspecting equipment for nicks, rips, tears and fit. Training was conducted for staff with COVID specific autopsy changes on a regular basis, amounting to approximately 12 hours per staff member over the course of 2020.

The enhanced PPE and added safety procedures were used for all autopsies regardless of knowledge of the presence or absence of the subject’s COVID-19 status.

### Autopsy procedural changes instituted for COVID-19

Staff performing autopsies were limited to two (reduced from an average of four) wherever possible. Instead of each leader having a dedicated helper, the leaders became each other’s helper.

Once the decedent was placed in the morgue, a precautionary sign was placed on the entry door stating, “Autopsy in Progress, Authorized Personnel Only, SARS-CoV-2 Awareness, Proper PPE Required.” This was the only entry point into the morgue. Once the procedure had begun, doors remained closed to retain negative air pressure in the autopsy suite. Before beginning cadaver dissection, a nasopharyngeal swab was collected for subsequent SARS-CoV-2 PCR testing done in a CLIA-approved laboratory with CDC-certified PCR tests. Following the nasal swab, the decedent’s face was draped with an alcohol-dampened cloth to reduce aerosols coming from the donor’s mouth and nose. The cadaver was then sprayed with a 70% alcohol solution to reduce micro-organism counts.

Autopsy procedures and sample collection were abbreviated to reduce the time of exposure while still providing essential sample collection for research. The scalp was resected and the skullcap removed with an oscillating electric saw, essentially as outlined in Sheaff and Hopster’s Postmortem Technique Handbook (2005), but using a shrouding technique similar to that published by MacArthur and Schneiderman (1987) to contain the generated aerosols at autopsy. A heavy-duty clear polyethylene bag was modified to create the shroud (HDX Commercial Drum Liner, 55-gallon, 1.7 mm thickness, clear, Model # HD55WC040C) by cutting through the closed end of the bag. This was placed over the decedent’s head and secured around the neck with a zip tie (cable tie, tie wrap), with the other side secured around the neck of the oscillating saw handle (Figure 1).

**Figure 1.**
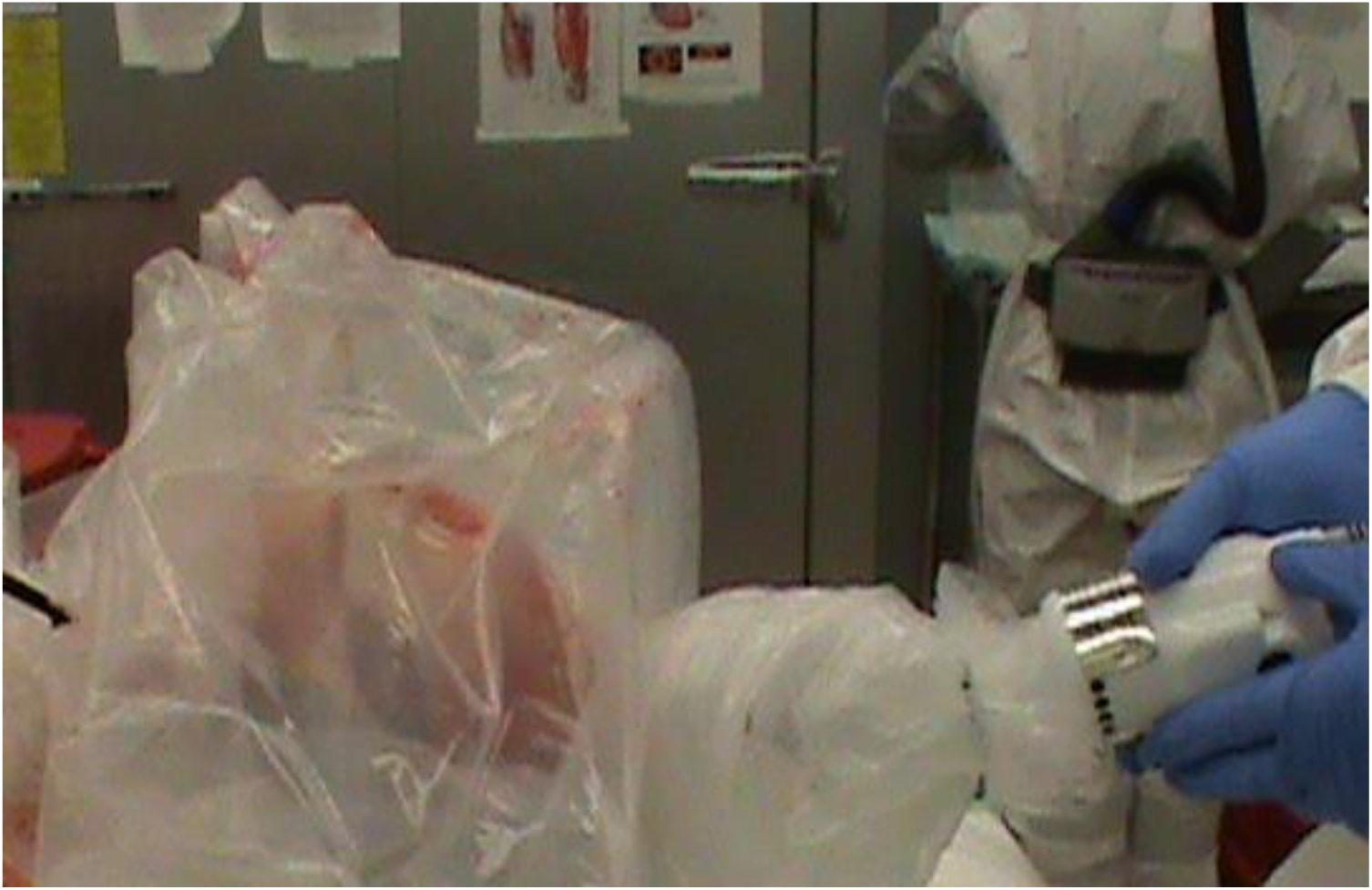
Image showing the use of a heavy-duty clear polyethylene shroud (made from commercial trash can liner) to limit aerosols while removing the skull cap.

The skull cap was then removed by gripping the saw handle through the shroud, being careful to not touch the blade to the bag or otherwise make any holes or tears. Once finished, the bag was carefully removed to a biohazard bin and brain removal completed.

Peripheral organ collection was limited to fewer organs, and the samples taken were generally a smaller sample size than taken before the COVID protocol was put in place. To allow a survey of SARS-CoV-2 RNA differential presence amongst multiple organs, the instruments used to collect the organs were exchanged with a fresh clean pair for each new organ to limit cross-contamination between organs. For the same reason, the prospector’s gloved hands were rinsed with sanitizer or 70% ethanol between each organ sampling. Only one technician at a time would be actively taking samples from the cadaver, to reduce the potential for unintentional blade injury. Following the recommendations of the University Hospital Basel, organs were no longer removed and weighed. (“Supplementary,” n.d.). Rather, organs were only qualitatively assessed in situ with the intention of reducing droplet spread and body fluid contact. As mentioned, the spinal cord was not collected in order to avoid aerosol generation with the bone saw and reduce exposure time in the autopsy room. Aorta sampling was limited to the thoracic part. After the rib cage was removed using gardening shears, towels were placed over the cut edges to prevent the risk of puncture or laceration. When slicing organs such as the spleen, kidney, and liver, a surgical towel was placed over the knife and organ while cutting to contain any excess fluid that might be expressed under pressure (Figure 2).

**Figure 2.**
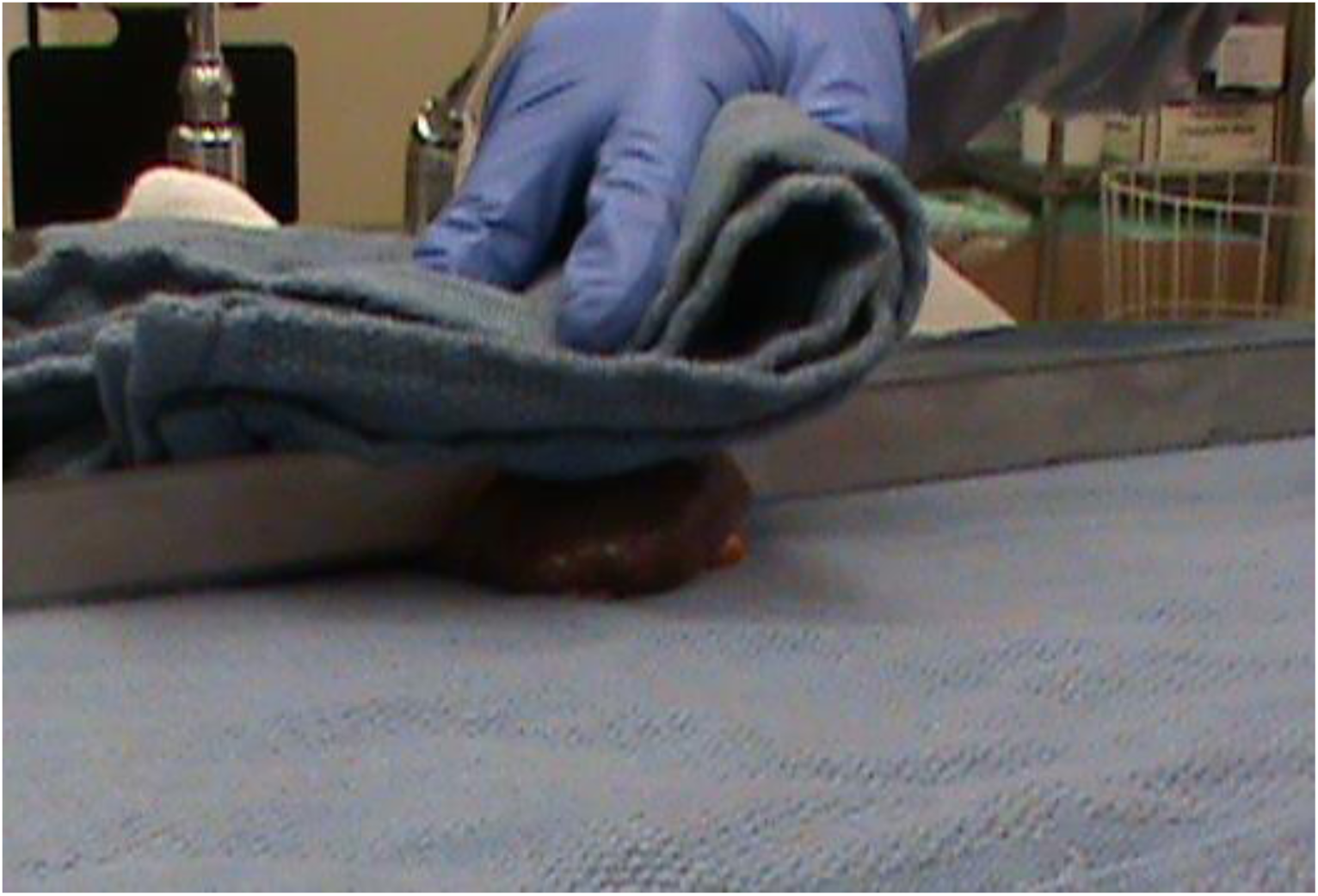
Image of the spleen being covered by a towel while sample is being cut with a knife.

Samples were divided in half with a knife or scalpel, with one half immediately immersed in 5% NBF and the other half immediately frozen on a slab of dry ice resting on a tin foil sheet.

Zip ties were used in place of string or suture material to contain luminal contents of the small and large intestines. Four zip ties were used to isolate and contain the contents of a short length of intestine (Figure 3).

**Figure 3.**
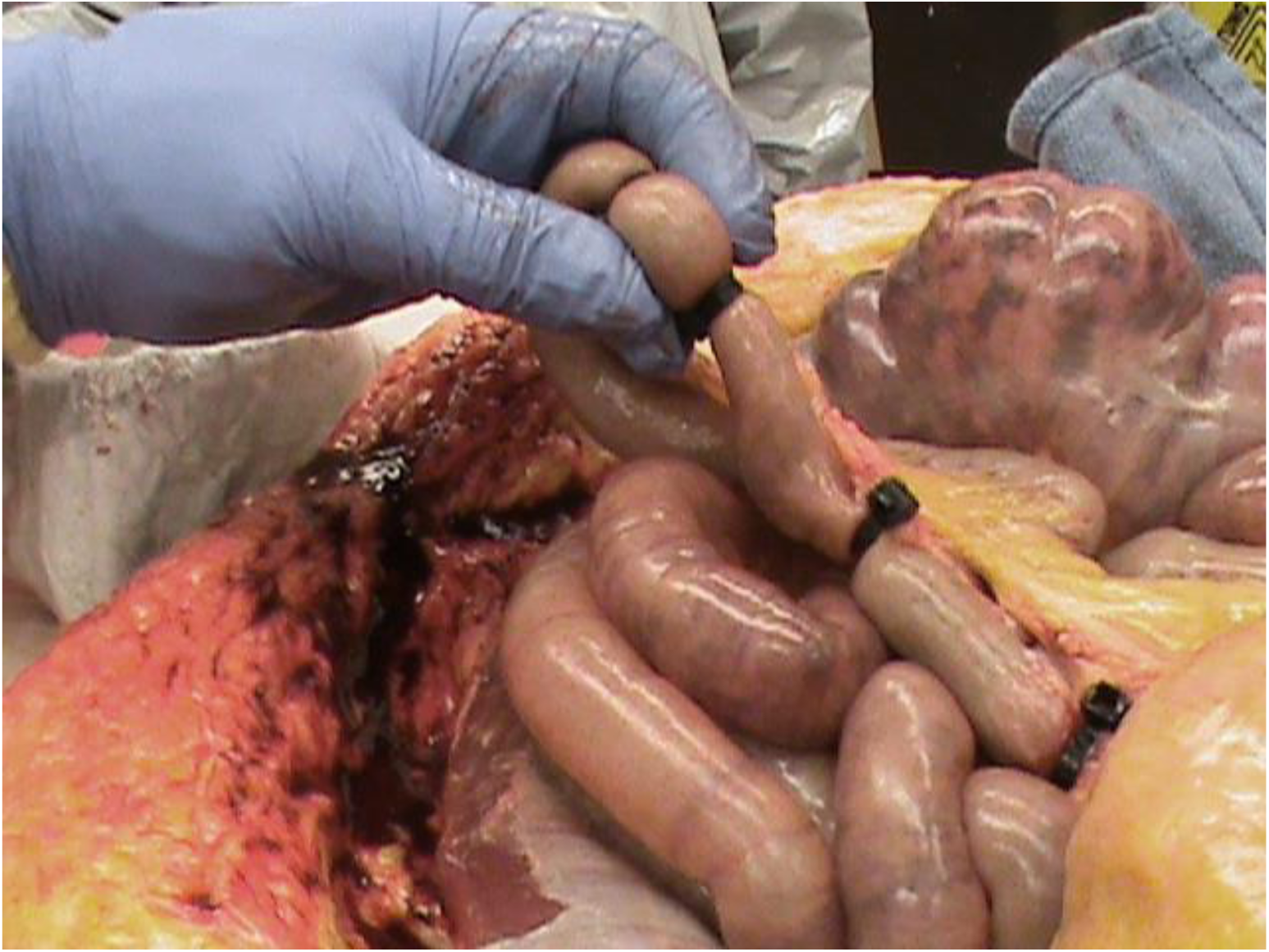
Image showing how zip ties were used to isolate and contain contents of sampled segments of intestine.

To shorten the protocol, some samples were no longer collected, including the duodenum, sigmoid colon, rectum, submandibular gland, larynx, esophagus, breast, cervix, prostate, gallbladder, skin, spinal cord, stomach, eyes, uterus, vagina, sciatic nerve, vagus nerve, psoas muscle, and mesentery.

However, additional samples hypothesized to be involved with COVID-19 were collected for the first time, including samples of the femoral nerve, quadriceps muscle, carotid artery, trachea, and aortic arch, as well as additional heart samples, including samples of the cardiac apex, interventricular septum, and left and right ventricular walls. The remainder of the heart was placed in 5% NBF. The entire lungs were collected, with slices of the superior-most upper lobes and inferior-most lower lobes frozen on slabs of dry ice while the remainder of the lungs bilaterally were placed in the same bucket with the heart to be fixed in 5% NBF. All areas of the respiratory tract were collected last to limit the exposure time to organs with presumably the most virus present. After one week fixation in the autopsy suite, the fixed samples and organs were further dissected into tissue cassettes for dehydration, paraffin infiltration and embedding.

Samples frozen in the autopsy room were transferred afterwards out of the autopsy suite to a sealed box in a -80 C freezer. All samples from individuals who died with active SARS-CoV-2 infections were stored at a dedicated -80 C ultra-low temperature freezer.

Once samples of all the organs were collected, the ribcage and remaining organs were returned to the cadaver and the thoracoabdominal incision was sewn up. Prior to departing the autopsy suite, the cadaver was cleaned with wet surgical towels, again sprayed with 70% ethanol and disinfectant, and transferred into a new vinyl body bag. The body was then transferred to the morgue walk-in refrigerator, adjacent to and directly connected to the autopsy suite, for short-term storage before being sent to the designated mortuary. In the autopsy suite, general cleaning was then performed, and all biohazard-contaminated materials were bagged and tied for disposal. The floor was mopped with disinfectant (e.g. Virex®), prior to leaving.

### Post autopsy procedures

After the autopsy was concluded, doffing was done according to procedures designed for Ebola and other viral hemorrhagic fevers (Centers for Disease Control and Prevention, 2020), in a two-stage area with HEPA air filtration and negative air flow. Autopsy team members assisted each other in doffing with supervision. As each layer of PPE was removed, team members sprayed the outermost layer with 70% ethanol. Care was taken to turn PPE inside out as it was removed to reduce the amount of exposed contaminated surfaces in the doffing area. All removed disposable PPE was placed directly into a biohazard bin. As soon as the PAPRs were removed, the team member put on an N95 mask. The PAPRs were sanitized with Virex® and left to dry. Once doffing was completed, hand hygiene was performed both with a hand sanitizer prior to leaving the doffing area and hand washing after leaving the doffing area. After doffing was complete and staff left the autopsy suite, it was undisturbed for a minimum of two hours to allow any generated aerosols to settle and for 54 air filtration cycles to be completed. The autopsy team would then put their used scrubs in a designated soiled container to be cleaned, and a fresh pair was put on. Between May 8, 2020, and February 28, 2022, all staff members underwent voluntary serial nasopharyngeal SARS-CoV-2 PCR diagnostic testing on a regular basis as well as immediately upon presentation of any COVID-19 symptoms. Staff testing positive remained at home until testing negative.

## Results

The BBDP handled 162 autopsies between the first reported case of COVID-19 in Arizona on January 26, 2020, through February 28, 2022. Of these, 36 individuals died with a positive nasopharyngeal PCR test for SARS-CoV-2, resulting in 94 individual exposures to the 19 staff members involved. During this period, there were 2 occurrences of a single staff member testing positive forCOVID-19 within seven days following their participation in an autopsy on an infected subject. However, one of these positive tests were associated with a known personal contact of that staff member with a SARS-CoV-2-positive individual in a different environmental setting (e.g. staff member’s family member or friend). No autopsy was followed by positive SARS-CoV-2 testing in more than one autopsy staff attending an autopsy. In comparison, the 19 autopsy staff members had 9 documented positive infections with SARS-CoV-2 over this same time period, without having participated in an autopsy on a SARS-CoV-2-positive subject within 7 days of their positive test (Table 2).

**Table 2.** Number of positive and negative Covid tests in correlation to autopsy. Fisher’s exact test p value is 0.6654, so there was not a significantly greater proportion of positive to negative tests within 7 days of a Covid autopsy as p>.05.

|  | Within 7 days of a Covid autopsy | Not related to a covid autopsy |
| --- | --- | --- |
| Number of positive tests in all team members | 2 | 9 |
| Number of negative tests in all team members | 92 | 573 |

These results suggest that our autopsy protocols radically reduced the risk of contracting COVID-19 as a result of performing autopsies on infected subjects. The institution of our enhanced autopsy protocols enabled us to conduct studies of COVID-19 that have been posted as 6 publications in preprint servers and peer-reviewed scientific journals (Serrano et al, 2022; Röltgen et al, 2022; Piras et al, 2021; Serrano et al, 2021; Tremblay et al, 2021; Beach et al 2021).

## Discussion

During the COVID-19 pandemic, pathologists, pathology technicians, histology technicians, and autopsy technicians worked to provide essential information about the way SARS-CoV-2 impacts the human body. In the early days of the pandemic, there were no experience-validated guidelines or standards for autopsies specific to individuals dying with COVID-19. At multiple institutions worldwide, the modifications made to autopsy protocols allowed COVID-19 to be studied while reducing the risk of being infected during autopsy. For example, our BBDP made changes to the autopsy protocol that allowed us to continue doing autopsies with minimized staff exposure and risk while contributing high quality information to the greater scientific community, both about SARS-CoV-2 and our original focus, neurodegenerative and cerebrovascular diseases causing dementia and parkinsonism. Thanks to autopsy studies like ours, invaluable information was published, informing the world medical community on the prevalence of SARS-CoV-2 viral brain invasion and the possible route of infection. We found that 38% of the cases that died with an active COVID-19 infection also had PCR-detectable SARS-CoV-2 RNA in their brain, but with a limited spread beyond the likely brain portal in the olfactory bulb (OB). In addition, we found that COVID-19-associated brain changes, including massive alterations of gene expression in the OB and amygdala, rather than being due to direct viral damage, may be more likely due to bloodborne immune mediators and transsynaptic gene expression changes arising from OB deafferentation (Serrano et al, 2022; Röltgen et al, 2022; Piras et al, 2021; Serrano et al, 2021; Tremblay et al, 2021; Beach et al 2021).

We did not suspend doing autopsies at any time, and our staff probably did not contract COVID-19 through autopsy exposures. We cannot really be certain, however, whether the 2 staff members testing positive within a week of an autopsy exposure were truly a result of that exposure versus exposure to an infected individual in the community. According to the Johns Hopkins Coronavirus Resource Center, between January 26, 2020, and February 25, 2022, Arizona had 1975252 confirmed COVID-19 cases, representing 26.8% of the state’s population. In comparison, during that same time period, 2 (10.5%) of our COVID-19-exposed autopsy staff at the time were infected, suggesting they may not have had a significantly increased risk as compared to the general population, especially considering the probable much higher rate of SARS-CoV-2 diagnostic testing done on autopsy staff.

## Conclusion

In response to the COVID-19 pandemic, the BBDP undertook significant changes in the engineering, environmental, and procedural ways autopsies were performed in order to continue our work. The revised protocol was shown to have a high probability of effective protection, as the incidence of SARS-CoV-2 infection amongst the BBDP was not significantly greater than that of the surrounding general population. Even though our autopsy protocol may have been more cautious than needed, with a pathogen of unknown transmissibility it is better to err on the stricter side in order to protect those performing this vital public service. We recommend the use of this protocol to other laboratories performing autopsies in future infectious disease outbreaks.

## Data Availability

All data produced in the present work are contained in the manuscript.

## Acknowledgements and Funding

We are all humbled by the volunteer individuals and their families that have selflessly consented to autopsy, allowing us to begin to understand the neurological consequences of COVID-19. This project was supported by a COVID-19 Supplement to a National Institute on Aging grant (3P30AG019610-20S1), submitted in response to a Notice of Special Interest (NOSI) issued by the National Institute on Aging (NOT-AG-20-022). Other support was provided by P30AG072980 to the Arizona Alzheimer’s Disease Research Center. Biospecimens from the Banner Sun Health Research Institute Brain and Body Donation Program, including those presented in this report, are available to qualified researchers upon request from https://www.brainandbodydonationregistration.org/.

